# Therapeutic targeting of alternative splicing caused by a lethal noncoding structural variant in X-linked dystonia parkinsonism

**DOI:** 10.1101/2025.10.10.25336355

**Authors:** Rachita Yadav, Christine A. Vaine, Aloysius Domingo, Siddharth Reed, Shivangi Shah, Dadi Gao, Kathryn O’Keefe, Monica Salani, John Lemanski, Riya Bhavsar, Moira A. McMahon, Michaela Jackson, Margo Courtney, Micaela G. Murcar, Cara Fernandez-Cerado, Gierold Paul A. Legarda, Michelle Sy, M. Salvie Velasco-Andrada, Edwin L. Muñoz, Mark Angelo C. Ang, Cid Czarina E. Diesta, Serkan Erdin, Ellen B. Penney, Laurie Ozelius, Nutan Sharma, C. Frank Bennett, D. Cristopher Bragg, Michael E. Talkowski

## Abstract

X-linked Dystonia-Parkinsonism (XDP) is a lethal adult-onset neurodegenerative disorder that exhibits features of dystonia and parkinsonism and is exclusively associated with a causal founder haplotype that is indigenous to the Philippines and affects Filipino males. Using patient-specific fibroblasts, neural stem cells (NSC), and other neuronal models, we discovered that cryptic alternative splicing caused by a novel SINE-VNTR-Alu (SVA) mobile element insertion into intron 32 of *TAF1* is a mechanistic hallmark of XDP. We leveraged postmortem brain samples from an XDP-specific brain bank to demonstrate that the molecular hallmarks of XDP observed in neural stem cells (NSCs) mirror abnormalities observed in brain tissues from affected patients. Based on these findings that patient-specific NSCs reproduce mechanistic signatures found in the brain, we sought to develop a bespoke precision therapeutic for XDP and evaluate its relative efficacy in ameliorating transcriptomic signatures in neuronal models. We first used CRISPR-based excision of the SVA and demonstrated ablation of all aberrant splicing and dysregulation of *TAF1* expression in NSCs across 30 independent clones. CRISPR-based correction of the XDP haplotype also restored the expression of 424 of 1,490 (30%) differentially expressed genes (DEGs) that were altered in XDP patient lines and greatly exceeded what would be expected by chance (p-value = 9.89e-87). While in vivo delivery of a gold standard CRISPR therapy is currently not feasible for XDP, we evaluated a tractable approach for Filipino patients by exploring the potential to modulate alternative splicing in XDP patients using antisense oligonucleotides (ASOs). To accomplish this, we developed a large-scale and well controlled functional genomics platform that screened eighty ASOs targeting intron 32 of XDP patients, followed by prioritization of lead ASOs based on attenuation of the alternative splicing signature. In transcriptome analyses across 1,550 libraries, we found that 8 of the 12 lead ASOs ameliorated the targeted XDP aberrant splicing. Moreover, we found that the two lead ASOs exhibited 38% and 43% rescue of XDP-specific DEGs that were also rescued by CRISPR excision of the SVA (enrichment p-values = 2.06e-13 and 2.27e-05, respectively). These rescues represented restoration of key molecular functions previously implicated in XDP, such as synaptic function, DNA-binding transcription factor activity, and gliogenesis. This study highlights a path to a potential targeted therapeutic for XDP and the capacity to exploit functional genomic signatures in patient-derived neural models to develop a scalable precision therapeutic platform for rare genetic disorders.

**Overview:** 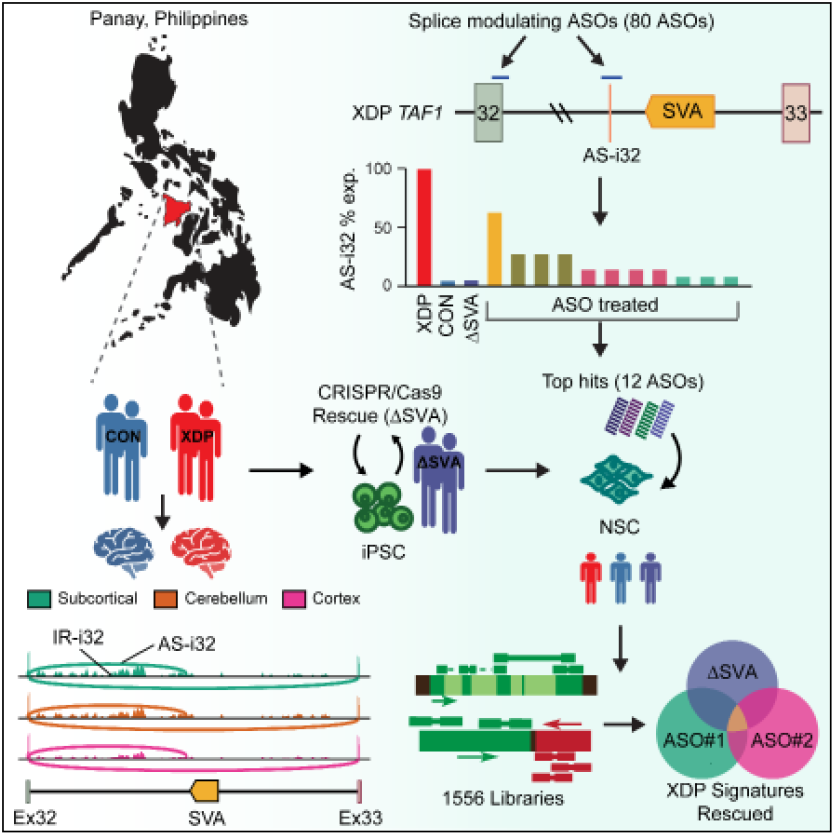

**Highlights:**

- A mobile element (SVA) insertion into TAF1 causes the severe neurogenerative disorder X-linked dystonia parkin-sonism (XDP) in Filipino males.
- The SVA mutation alters splicing of TAF1 in neuronal models and postmortem brains.
- CRISPR/Cas9 excision of the SVA rescues XDP alternative splicing and genome-wide transcriptomic signatures in neuronal models.
- ASOs precisely targeting the SVA insertion also rescue XDP molecular signatures.
- ASO based precision therapies rescue transcriptome-wide expression signatures that are shared with the CRISPR editing of XDP-specific SVA.

**In brief:** This work establishes a precision therapeutic framework for rare neurodegenerative disorders by genome editing and antisense oligonucleotide interventions in patient-derived neural models.

## INTRODUCTION

There are 250-450 million people worldwide that are afflicted with a rare disease at any point in time, with consequent morbidity, reduced quality of life, and early death for approximately 5.9% of the global human population.^1^ These diseases are predominantly genetic in etiology, and neurological changes represent the largest clinical manifestations.^2–4^ While there is still a significant unmet need for approved therapies for these rare genetic disorders, there are also exemplars of successfully identifying targets for genetic mutation-guided precision therapeutics, even in the most devastating conditions.^5–9^

X-linked dystonia-parkinsonism (XDP) is a severe neurodegenerative disorder for which no disease-modifying therapy exists to date. Treatment of symptoms is often insufficient in reducing pain, disability, and the burden of disease in afflicted individuals. This movement disorder is unique in its clinical presentation as well as its global distribution as it is caused by a founder effect and is most prevalent in Panay Island, Philippines.^10^ Neurological symptoms vary among individuals but generally include adult-onset dystonic posturing and loss of voluntary muscle control, followed by severe posture, gait, speech, and swallowing difficulties as the disease progresses.^11–14^ Neuroimaging and neuropathologic studies converge on involvement of subcortical brain structures, significant loss of neurons in the striatum, and presumably secondary pathologies in other brain regions.^15–18^ All individuals with XDP harbor a 2.6-kb SINE-VNTR-Alu (SVA) retrotransposon insertion in intron 32 of *TAF1*; the haplotype bearing this genetic variant has never been observed in other populations.^19–22^ Further, the length of a hexameric repeat within this SVA displays a surprisingly strong association with age-at-onset of XDP^23^ as well as clinical symptoms^24^. Tissue-specific hexameric repeat-length dependent somatic instability and mosaicism have been observed,^25–27^ and modified by alleles in DNA repair genes.^28,29^

Recent genome-wide profiling of structural variants (SVs) has clarified the frequency, diversity, complexity, and selective pressures against SVs - including large copy number variants, SVAs, and other transposable elements - in human genomes.^30–35^ Functional studies demonstrate that SVs impact local and global transcription regulation in neural tissue^36–39^ and are themselves transcriptionally active.^40^ SVA insertions can alter the exon and transcript structure of host genes via exon skipping or trapping mechanisms,^41,42^ and/or by activating cryptic alternative splice sites.^43^ Moreover, SVAs have been shown to alter epigenetic marks and modify interaction between promoter regions and transcription regulatory elements.^44^ We previously showed that the disease-specific SVA in XDP is associated with local effects, including aberrant alternative splicing in the vicinity of the SVA, partial intron 32 retention, and dysregulation of *TAF1* expression.^22^ Mechanistically, these transcriptional alterations are thought to be due to the SVA directly repressing *TAF1* promoter activity^45^, altered epigenetic silencing of the SVA during development or with aging^46,47^, and/ or by transcriptional interference by G-quadruplex structures and R-loops arising in GC-rich regions of the SVA.^27,48^ Irrespective of mechanism, there is an intimate causal link between the disease-specific SVA in XDP and local splicing and expression hallmarks in *TAF1*, as evidenced by effective correction of these transcription abnormalities via CRISPR-excision of the SVA in cell lines.^22,49^

While *in vivo* CRISPR editing has recently been demonstrated to be feasible,^5^ there are a myriad of challenges inherent in payload delivery and efficiency of CRISPR excision in the central nervous system, making this method currently less attractive as a therapeutic option in neurodegenerative disease. In contrast, modulatory effects at the level of RNA can be effectuated by antisense nucleotides (ASOs). ASOs are synthetic nucleic acid fragments that bind via Watson-Crick base-pairing to a target sequence. There are a variety of mechanisms in which they can modulate the target RNA, including RNA degradation via recruitment of endogenous ribonuclease H (RNase H),^50^ alterations in RNA stability via chemical modifications,^51^ and modulation of splicing by directly binding to exon/intron junctions, cryptic splice sites, or enhancer/repressor regions.^43,52^ Currently, there are approved ASO-based treatments for spinal muscular atrophy and Duchenne muscular dystrophy.^53–57^ For other neurodegenerative diseases such as Huntington’s disease (HD), the spinocerebellar ataxias, amyotrophic lateral sclerosis (ALS), multiple system atrophy, prion disease, and Parkinson’s disease (PD), ASO-based therapies are in various stages of development, or approved in the case of tofersen^58^. Given the high specificity with which ASOs can be designed to access and modulate their targets, they can be appropriate tools for diseases with sufficient knowledge of the genetic architecture and pathologic expression signature.

Here, we sought to develop a bespoke precision therapy for XDP, leveraging our previous discoveries of a causal noncoding SVA in XDP and the expression hallmarks of aberrant splicing and dysregulation of *TAF1* function. We first demonstrate robust and reproducible correction of XDP-associated splicing and expression deficits across a large set of CRISPR-edited neural stem cell (NSC) clones from XDP patients. We then utilized postmortem tissues from an XDP brain bank^59^ to show that the signatures we discovered in patient-specific cell lines are ubiquitous across affected neuronal tissues, suggesting that neuronal models and transcriptional signatures offered strong platforms for therapeutic development. We screened 80 ASOs targeting alternative splicing in XDP and carried 12 lead ASOs for comprehensive functional genomic evaluation. We observed rescue of splicing and altered gene and pathway transcriptional signatures in XDP from the lead ASOs, including two consistently reproducible compounds, demonstrating that ASO-based therapeutics offer a viable precision medicine for XDP. Our analyses further suggest that patient–specific neuronal models of disease and transcriptomic profiling offer new opportunities for therapeutic development in rare diseases.

## RESULTS

### A cryptic alternative splice junction in TAF1 intron 32 is ubiquitous in XDP patient-derived tissues and cells

We previously established that a 2,627-bp SVA insertion in intron 32 of the *TAF1* gene underlies the genetic cause of XDP (**Figure 1A**). We performed *de novo* assembly of the genome and transcriptome of XDP patients at the time of that study and did not observe transcripts directly emanating from the SVA; instead, we discovered a novel cryptic splice junction (AS-i32) within 700-bp of the insertion site of the SVA insertion that is unique to XDP patients^22^ (**Figure 1A**). We thus developed an expanded collection of induced pluripotent stem cell (iPSC)-derived NSC lines from patients and controls to interrogate AS-i32 (**Supplementary Table S1**). We queried all expressed splice junctions from the TAF1 locus, including extremely rare junctions using targeted deep transcript sequencing (RNA-CapSeq), and observed consistent expression of AS-i32 in all 34 XDP NSC lines tested (average normalized counts = 425.4). In contrast, in NSCs from controls, we noted ∼185-fold reduction of this splice junction (mean normalized counts = 2.3 with median = 0), confirming the specificity of this cryptic alternative splice junction in NSCs from XDP patients (**Figure 1C, Supplementary Table S2**).

**Figure 1.**
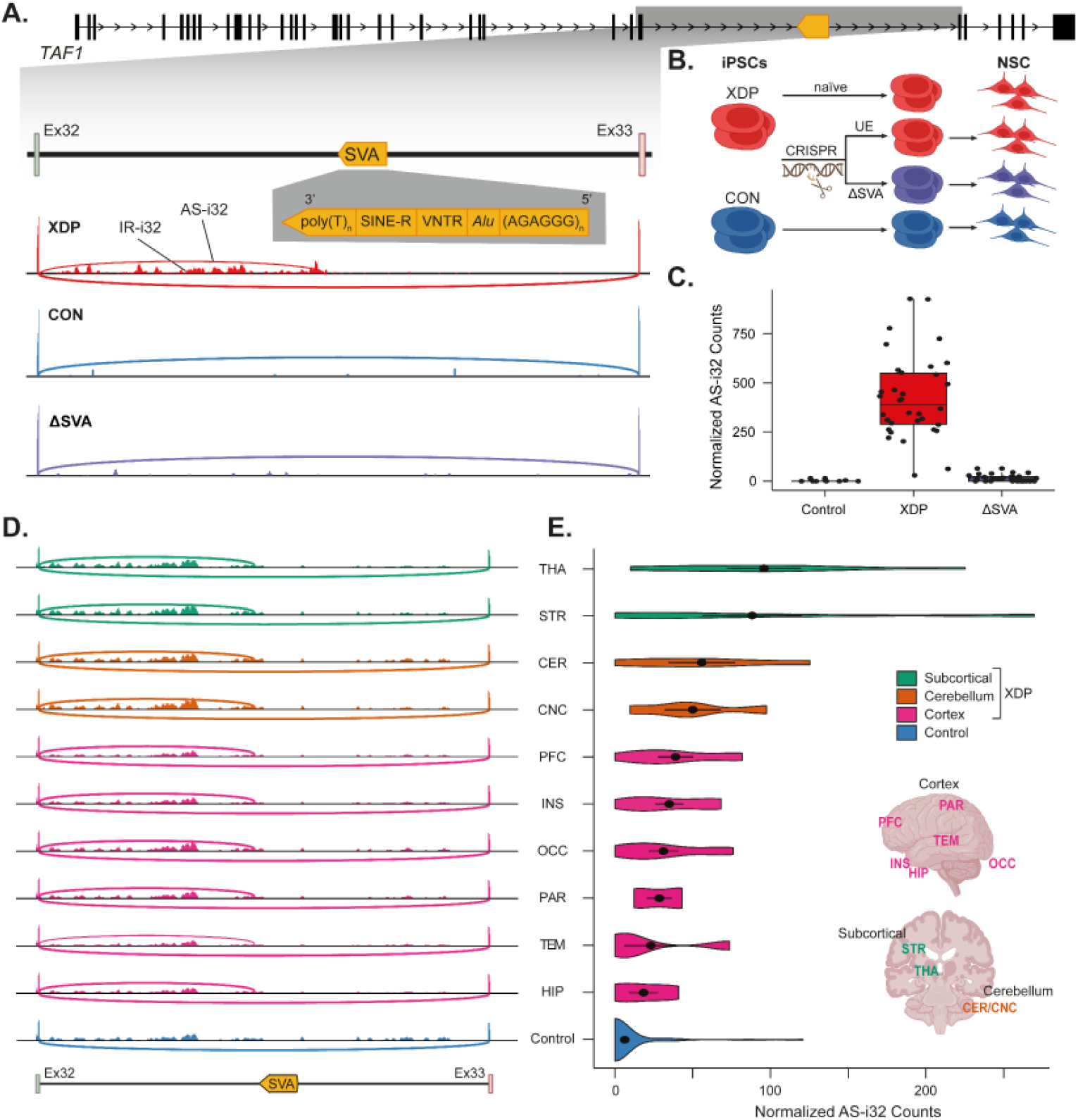
Observation of the cryptic alternative splice junction in *TAF1* intron 32 in post-mortem XDP brains. **A)** XDP-associated *TAF1* gene architecture with SVA insertion in intron 32. XDP-associated alternate spliced transcript (AS-i32) and intron retention (IR-i32) in *TAF1* intron 32 in NSC from XDP patients and controls. **B)** Experimental design of the SVA CRISPR editing (ΔSVA) in patient-derived iPSCs, followed by differentiation into NSCs, along with naive iPSCs, CRISPR-exposed but unedited (UE), and control lines. **C)** Normalized expression of AS-i32 in NSC from XDP patients and controls and change in AS-i32 with SVA excision by CRISPR (ΔSVA). Each point represents one individual. **D)** AS-i32 and IR-i32 signatures in NSC are mirrored in postmortem brains from XDP. PFC - prefrontal cortex, PAR - parietal cortex, TEM - temporal cortex, INS - insular cortex, HIP - hippocampus, OCC - occipital cortex, STR - striatum, THA - thalamus, CER - cerebellar cortex, CNC - cerebellar nuclei. **E)** Variation in *TAF1* AS-i32 across different regions in XDP and control postmortem brain samples.

To further explore the relationship between the disease-specific SVA in intron 32 and the expression of AS-i32 in XDP cells, we used genome editing to excise the SVA from XDP lines while keeping the rest of the genome intact (**Figure 1B**). We performed CRISPR/Cas9-based excision in iPSCs derived from six XDP probands and differentiated them to NSCs, including up to five successfully edited clones (ΔSVA) and up to five matching unedited-XDP clones from each XDP parent line, for a total of 48 NSC lines. In 30 ΔSVA NSC lines, we observed a 98.4% reduction of AS-i32 compared to unedited-XDP (mean fold-change [fc] = 0.016, 95% confidence interval [0.009-0.030]), bringing AS-i32 expression in these lines approximately to the level in unaffected controls and representing a near complete rescue (**Figure 1C**). In contrast, CRISPR-targeted but unedited XDP lines retained expression of AS-i32, reduced *TAF1* expression, and overall transcriptomic profiles that recapitulated those observed in untargeted XDP NSCs (**Supplementary Figure S1A-D**). These results demonstrate a clear amelioration of aberrant transcriptional signatures via *in vitro* CRISPR manipulation of the causal SVA mutation in XDP.

To determine whether this expression signature in developmental cell lines proxies XDP splicing abnormalities in mature neurons in patients, we obtained 19 XDP postmortem brain samples from a disease-specific brain bank created specifically for these studies^59^ and investigated the presence of AS-i32 in brain tissue (**Figure 1D**). Matched brain regions from 9 individuals without XDP phenotype were obtained from the NIH NeuroBioBank to serve as controls. The splicing pattern identified in XDP *in vitro* was also observed in postmortem brain (n regions = 10, **Figure 1E**), albeit at lower expression levels in comparison to AS-i32 expression in actively cycling NSCs (**Supplementary Table S2**). Notably, in brain regions that have been previously implicated in XDP such as the striatum, thalamus, or cerebellum,^15–17,60^ we observed higher AS-i32 expression in comparison to cortical regions (highest AS-i32 in thalamus, average normalized counts = 95.8; striatum, average normalized counts = 88.3; cerebellar cortex and dentate nucleus, average normalized counts = 55.9 and 50.0, respectively) (**Figure 1E**). Consistent with our findings in cell models, AS-i32 was not reproducibly observed in brain tissue from controls (median normalized counts in controls = 0, mean = 6.3 across eight brain regions).

### ASOs successfully repress XDP aberrant splicing in neuronal models

Our results in NSCs and postmortem brain confirmed that the splicing defect AS-i32 is a consistent expression marker of XDP, rendering it an attractive target for ASO modulation. Guided by the successes of multiple clinical ASOs targeting pre-mRNA splicing,^61–64^ we designed a set of ASOs targeting sequences surrounding the AS-i32 splice junction in intron 32 in *TAF1* (**Figure 2A**). A total of 80 splice-modulating ASOs that tiled a 200-bp region up/downstream of the AS-i32 splice junction were tested in an initial screen using targeted qPCR assays to identify positive hits based on successful modulation of AS-i32 message levels in transfected fibroblasts. Of the 80 ASOs, 35 showed >80% knockdown of AS-i32 in these cells (**Figure 2B, Supplementary Table S3**). Of the top 12 potential ASOs, 11 mapped to a region 7-81 bp upstream of the AS-i32 splice junction and one mapped distal to AS-i32.

**Figure 2.**
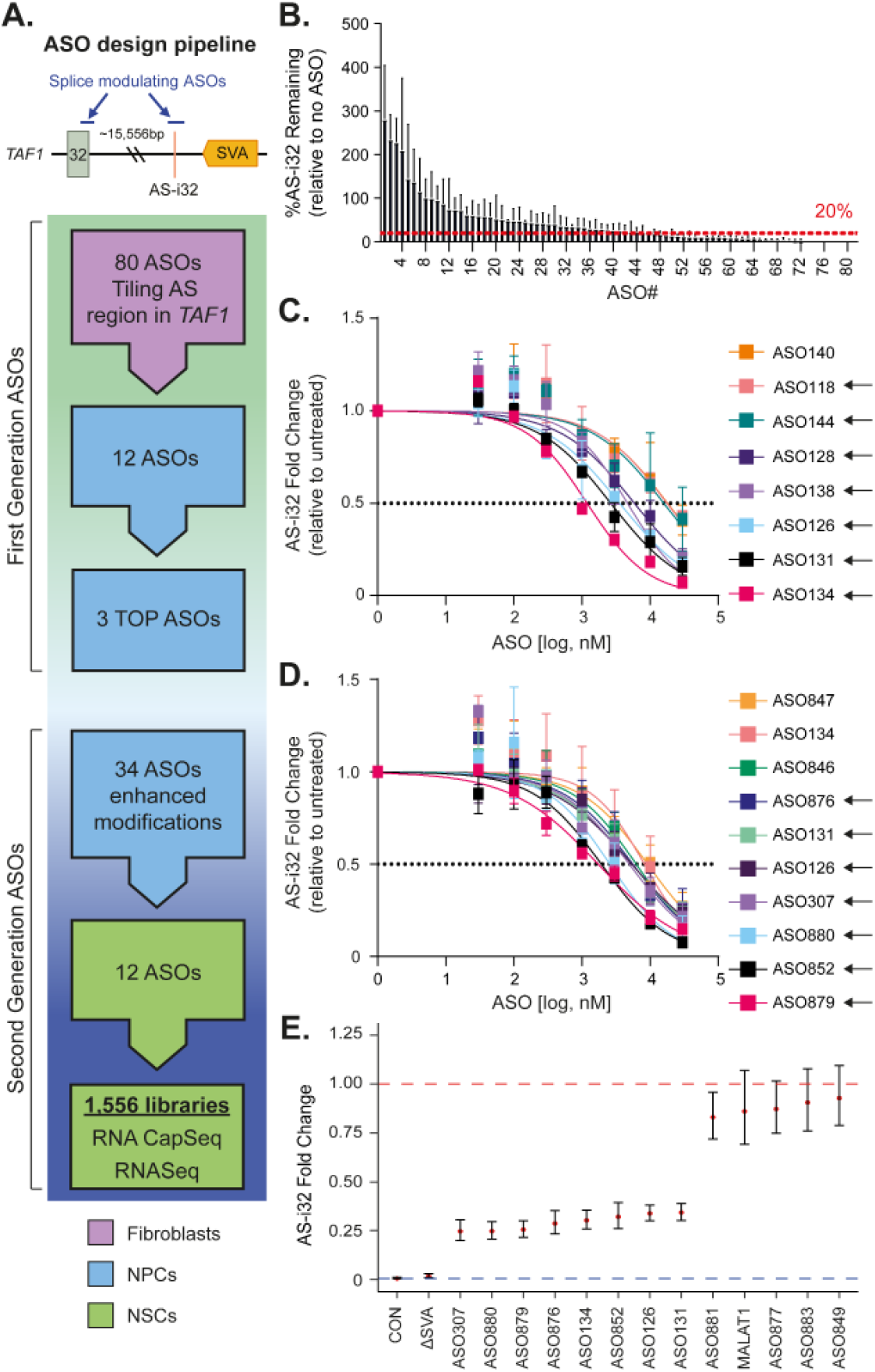
Design and testing of ASOs against the XDP-specific cryptic alternative splice junction in *TAF1* intron 32 (AS-i32). **A)** ASO design pipeline. **B)** Initial AS-i32 target screening of potential ASOs in transfected fibroblasts. **C, D)** Dose-response curve analysis of original **(C)** and second-generation **(D)** ASOs in NPCs. ASOs were added to the culture media of two **(C)** or three **(D)** XDP NPC lines at the given concentrations. AS-i32 expression levels were assayed after 3 days. To calculate EC50s, fit curves were calculated using nonlinear fit (log(inhibitor) vs. response - variable slope) and constraining the top and bottom values at 1 and 0, respectively. The dashed line indicates 50% expression. Data expressed as mean ± SEM. **E)** Rescue of AS-i32 levels in NSC by CRISPR and ASOs. ASOs (5 μM) were added to NSC culture media for 7 days. Comparison against untreated XDP (red line) and control AS-i32 profile (blue line).

To determine targeting efficiency AS-i32, we performed dose response experiments using the 12 ASOs in two XDP neural progenitor cell (NPC) lines, as NPCs express AS-i32 at a higher level in comparison to fibroblast lines and functionally uptake ASOs passively from the cell culture media. AS-i32 levels were assayed via qPCR three days after the addition of ASOs in culture media^22^. Of the 12 ASOs, eight exhibited dose-dependent AS-i32 reduction, where >50% knockdown was observed at the highest concentration (**Figure 2C, Supplementary Table S4**). A rank order based on EC^50^ was generated, with the top three ASOs exhibiting EC^50^s of less than 4 μM. We then designed 34 second-generation ASOs using sequences derived from the top three ASOs (ASO126, ASO131, ASO134) and applied additional design features and chemical modifications to optimize stability, uptake, and target engagement (**Figure 2A, Table 1**). Following a 3-day treatment in three NPC lines, seven out of 34 ASOs, along with the three parent ASOs, exhibited a dose-dependent AS-i32 reduction (**Figure 2D, Supplementary Table S5**). Encouraged by this data, we treated a large NPC inventory composed of 9 XDP, 2 ΔSVA, and 5 control clones with all 37 ASOs at the optimized concentration of 5 μM and determined AS-i32 expression via RNA-CapSeq (total 669 libraries). We identified 12 ASOs that repress AS-i32 by up to 85.5% in this experiment (**Supplementary Figure S2**) (minimal mean fc = 0.155 [0.060-0.400] in ASO880), corroborating the top hits from the EC_50_ analyses.

**Table 1.** List of second-generation ASOs, grouped by parent ASO (126, 131, or 134).

| Family |  |  |
| --- | --- | --- |
| ASO 126 | ASO 131 | ASO 134 |
| 126 | 131 | 134 |
| 307 | 843 | 851 |
| 837 | 844 | 852 |
| 838 | 845 | 853 |
| 839 | 846 | 854 |
| 840 | 847 | 879 |
| 841 | 848 | 880 |
| 875 | 849 | 881 |
|  | 876 | 882 |
|  | 877 | 883 |
|  | 878 | 890 |
|  | 889 | 891 |
|  | 070 | 073 |
|  | 071 | 074 |

Given our discovery that excision of the SVA via CRISPR reproducibly ablates AS-i32 in XDP NSCs, this intervention serves as a benchmark comparator for ASO modulation therapies. In our final screen, we treated 68 NSC clones (9 *naïve* XDP, 21 unedited XDP, 30 ΔSVA, 8 controls) with the 12 ASOs from the dose response for seven days, including the three parent ASOs, and an uptake control ASO targeting *MALAT1*. We performed RNA-CapSeq and investigated AS-i32 expression in a total of 1,556 libraries (**Figure 2E**). Eight of the 12 ASOs significantly reduced AS-i32 in treated XDP lines (minimal mean fc = 0.246 [0.200-0.304] in ASO307) (**Figure 2E, Supplementary Table S2**), whereas the non-targeting negative control ASO-*MALAT1* did not have an effect. The splice modulating ASOs achieved up to 75% rescue of the aberrant AS-i32 expression compared to untreated XDP lines in NSCs.

### Rescue of TAF1 expression signatures by splice-modulating ASOs

We previously established that the aberrant splicing signature of the XDP-specific SVA involves AS-i32 as well as partial intron retention (IR) of the proximal segment of *TAF1* intron 32 (IR-i32),^22^ and we find that this signature is reproduced in postmortem XDP brains in this study. Intron 32 was the only *TAF1* intron where IR differed between XDP vs. controls (p-value < 0.10) in multiple brain regions (cerebellum, thalamus, striatum, and insular cortex) (**Supplementary Figure S3**). In contrast, in the occipital cortex, a region where no obvious XDP neuropathology has been described, no significant IR-i32 was observed in XDP compared to controls in these samples (**Figure 3A, Supplementary Table S5**).

**Figure 3.**
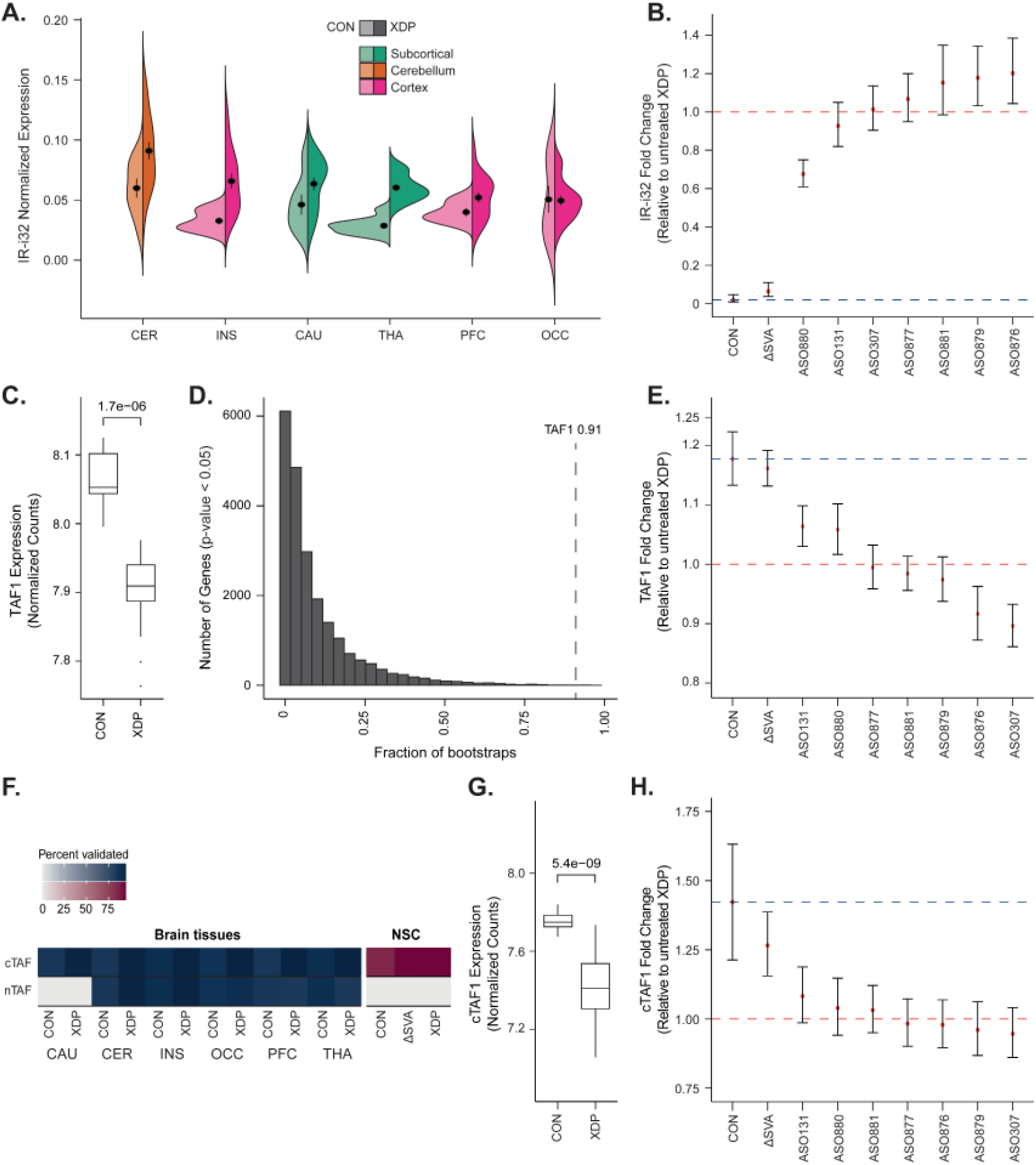
Establishment of TAF1 expression signatures and their rescue by splice-modulating ASOs. **A)** IR-i32 in postmortem brain regions from XDP and controls. **B)** Rescue of IR-i32 in NSC by CRISPR and ASOs. Comparison against untreated XDP (red line) and control IR-i32 profile (blue line). **C)** *TAF1* expression in NSCs. **D)** Distribution of frequency in the bootstrap data of genes that are down-regulated (LFC < 0) in XDP at p-value < 0.05. The dotted line indicates the frequency of *TAF1*. **E)** Rescue of overall *TAF1* expression in NSC by CRISPR and ASOs. Comparison against untreated XDP (red line) and control *TAF1* profile (blue line). **F)** Abundance of canonical (cTAF1) and neuronal (nTAF1) transcript variants across different brain regions, and their expression in NSCs. **G)** cTAF1 expression in XDP and CON NSCs. **H)** Rescue of canonical *TAF1* expression in NSC by CRISPR and ASOs. Comparison against untreated XDP (red line) and control *TAF1* profile (blue line).

In NSCs, IR was signifi cantly increased in XDP compared to control lines (mean fc = 50.7 [21.6-118.9]) and also rescued via excision of the SVA (ΔSVA NSCs: mean fc = 0.064 [0.037-0.109]) (**Figure 3B, Supplementary Table S6**). We thus anticipated that the ASOs used to repress AS-i32 would also lead to a reduction of local IR in NSCs. To investigate this, we generated RNASeq libraries from NSCs treated with seven ASOs prioritized by their successful manipulation of AS-i32 and identified an AS-i32-targeting ASO (ASO880) that also signifi cantly reduced IR-i32 expression by 32.4% compared to the XDP baseline (mean fc = 0.676 [0.610-0.750]) (**Figure 3B, Supplementary Table S6**).

*TAF1* downregulation was also observed in XDP cell models and tissues, a result that was most prominent in actively dividing cells by comparison to terminally differentiated neurons or brain tissues.^22,65–67^ Consistent with *TAF1* being an essential gene in males, and the predicted impact of a noncoding SV underlying a late-onset neurodegenerative condition, the magnitude of the regulatory impact of the SVA on *TAF1* expression is reproducibly subtle (19% in Aneichyk et al.^22^ and 18% in this study, mean fc = 0.849 [0.810-0.880] **Figure 3C**). Given the fundamental challenges with reproducibility in functional genomics studies using expression as a biomarker, we empirically assessed the expected variance and magnitude of effect of our expression profi ling across cellular models. We leveraged the power of the 1,556 libraries in this study to examine the reproducibility of *TAF1* discovery as a differentially expressed gene (DEG) in NSCs across thousands of Wald tests using randomly generated subsets of 6 XDP RNASeq profiles and 6 controls. In the 52,800 bootstraps performed, despite its relatively subtle perturbation, we observed that *TAF1* was significantly downregulated in XDP cases compared to controls in 98% of bootstraps. We further determined that *TAF1* was a statistically significant DEG in 91% of bootstraps at a probability threshold of p-value < 0.05 and in 85% of bootstraps at FDR < 0.10, suggesting good calibration of our statistical models against discovery power given the expression of abundance of *TAF1* in experiments comprised of 12 samples (6 controls, 6 XDP lines; **Figure 3D, Supplementary Figure S4, Supplementary Results and Supplementary Table S7**).

Following the quantification of the reproducibility of these results using *TAF1* expression as a target, we compared RNA-Seq profiles of untreated XDP lines with those of patient-matched ΔSVA-or ASO-treated NSCs to determine the magnitude of *TAF1* rescue. We found that CRISPR manipulation increased *TAF1* expression by 16%, thus representing a near complete rescue compared to controls (mean fc = 1.161 [1.132-1.192]). A subset of ASOs also rescued expression of *TAF1* in NSCs by up to 33% of control levels and 6% overall, including one the two lead compounds from the AS-i32 screening (ASO131; mean fc = 1.064, [1.030-1.099]) and both AS-i32 and IR-i32 screening (ASO880; mean fc = 1.058 [1.017-1.102]) (**Figure 3E, Supplementary Table S8**).

### Assembling TAF1 isoform expression in XDP brains

*TAF1* undergoes extensive alternative splicing, and brain-specific transcript variants have been described previously.^20,68,69^ To understand this diversity, we performed assembly and quantification of transcript variants using a combination of RNACapSeq and RNASeq in 6 postmortem brain regions from XDP and controls. We probed these assemblies for the canonical *TAF1* transcript (cTAF1/TAF1-008: ENST00000276072) as well as for neuron-specific transcripts that contain an alternative exon 34’ (nTAF1) that have been previously described with relevance to XDP.^70–73^ We found that cTAF1 was the major transcript across brain regions profiled, representing a median of 42.4% of overall *TAF1* expression across samples (**Supplementary Table S9**). In contrast, nTAF1 was inconsistently detected across samples and observed in less than 40% of caudate samples from both cases and controls. With the near-complete transcript structures of *TAF1* catalogued from these brain tissues, we were able to interrogate transcript-specific alteration in XDP brains and compare them to NSCs. We observed a reduction of cTAF1 specifically in the caudate of XDP patients (p-value = 0.03), a finding recapitulated in NSCs (p=5.45e-09, **Figure 3G**). As in the postmortem brains, cTAF1 was the predominant transcript in NSCs, contributing a median of 62.6% of overall *TAF1* expression (**Supplementary Figure S5A-B**). Analysis of *TAF1* isoforms showed a preferential rescue of cTAF1 expression in ΔSVA lines (up to 61% of control levels, or 26% overall; mean fc = 1.265 [1.154-1.386]), ASO131 (19% of controls; mean fc = 1.081 [0.985-1.187]) and ASO880 (7% of controls; mean fc = 1.039 [0.940-1.147]) (**Figure 3H**). These analyses indicate that therapeutic manipulation via CRISPR likely exerts the strongest effects by modulating the cTAF1 transcript of *TAF1*.

### Rescue of transcriptional dysregulation in XDP-associated pathways

TAF1 is a ubiquitously expressed protein; it is a transcription factor (TF) that is the largest subunit of the TFIID complex, and the gene is highly constrained in males.^74^ We thus hypothesized that subtle regulatory perturbations in *TAF1* (18% on average in XDP neural cells) could result in global gene dysregulation of relevant genes and pathways, and rescue of these pathways could provide insights into therapeutic development and efficacy. After robustly demonstrating through bootstrapping that no significant differences exist in gene expression profi les between XDP NSCs exposed and unexposed to CRISPR targeting, we combined these samples to build a large compendium of XDP NSCs harboring the causal SVA mutation and compared them to unaff ected controls (**Supplementary Figure S1A-C, Supplementary Tables S10-11**). At our well-calibrated FDR<0.10 threshold, the comparison of XDP versus control transcriptomes revealed 1,490 DEGs (590 upregulated and 900 downregulated DEGs) that we refer to herein as ‘XDP-signature genes’ (**Supplementary Figure S6, Supplementary Table S12**). These XDP-signature genes were functionally enriched for multiple ontology terms related to gliogenesis, cell adhesion, synaptic membrane, regulation of transcriptional activity, peptidyl serine phosphorylation, cell and receptor signaling, and neural cell differentiation (Bonferroni-corrected p-value <0.05, **Figure 5A**). Relevant to the phenotypes observed in XDP, the most significantly enriched Human Phenotype Ontology (HPO) term among XDP-signature genes was ‘poor fine motor coordination’ (p-value = 0.006; **Supplementary Table S13**).

We next assessed the impact of therapeutic manipulation via CRISPR editing (ΔSVA) and ASO treatment on XDP-signature genes and the potential for amelioration to converge on a critical subset of rescued genes and pathways. We discovered that 456 XDP-signature genes (31%, including *TAF1*, **Figure 4A**) were no longer differentially expressed in ΔSVA lines compared to controls (‘non-DEGs’; p-value > 0.05), thus reflecting transcriptome-wide rescue. Analyses of gene expression patterns emphasized that these results displayed extraordinarily high specificity; 93% of genes displayed reversal of their expression in XDP (e.g. down-regulated genes in XDP were upregulated following CRISPR; N=424/456, Fisher’s exact test p-value= 9.89e-87, **Figure 4B-C**). These findings suggest that CRISPR intervention was strongly associated with attenuating global dysregulation in gene expression associated with XDP beyond *TAF1* itself. Rescued gene-sets were enriched for functions associated with XDP, including regulation of TF activity via DNA binding motif, neural development (gliogenesis, neural differentiation), synaptic activity (synaptic membrane, glutamatergic and GABAergic synapses), protein modification (ie, phosphorylation), and signal transduction (**Supplementary Table S14**).

**Figure 4.**
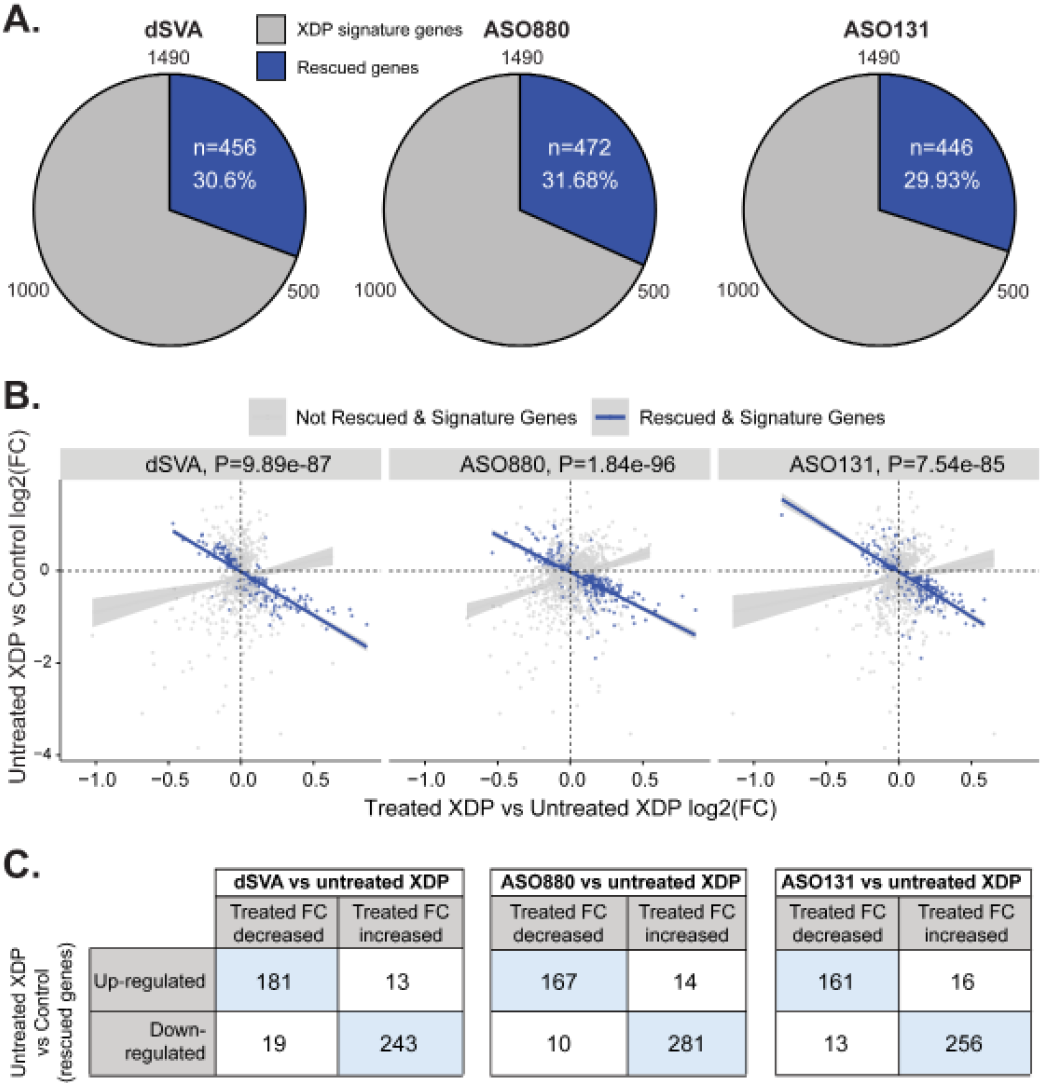
Directional rescue of XDP-associated signature genes by dSVA and top ASOs. **A)** Comparable rescue of XDP signature genes by genetic manipulation and ASOs. **B)** Directional reversal of XDP-signature genes after CRISPR and ASO treatment. P-value is based on Fisher’s exact test. **C)** 2X2 table showing reversal of directionality by comparing fold changes of rescued XDP-signature gene and their expression post-treatment by CRISPR and ASOs.

We then performed transcriptome-wide rescue analyses for each of the six ASOs that effectively ameliorated AS-i32 (ASO131, ASO307, ASO876, ASO879, ASO880, and ASO881). Sequence analysis of these ASOs revealed that they collectively target a 34-bp region surrounding the AS-i32 junction. Using the same analytic design as the CRISPR-based treatment, we found that the two lead ASOs both rescued global XDP-signature genes to a comparable extent as CRISPR; ASO131 rescued 446 DEGs (30%, including *TAF1*) while ASO880 rescued 472 DEGs (32%) (**Figure 4A**). When compared against the 456 genes rescued by CRISPR treatment, ASO131 and ASO880 rescued 197 (43%, enrichment p-value = 2.06e-13) and 172 (38%, enrichment p-value=2.27e-05) DEGs, respectively. Consistent with the observations in ΔSVA rescue, ASO rescued genes also exhibit greater directionality relative to the XDP-signature gene set. In the treated versus untreated comparison, a strong positive correlation would be expected if the drug had no biological effect. The observed deviation from this expectation indicates that the drug meaningfully perturbs the regulation of these genes, supporting the interpretation that the rescue process identifies genes with distinct, rather than random, responses. This directional bias, therefore, provides evidence for a genuine and targeted drug effect (**Figure 4B-C**).

Rescued genes again converged on XDP-associated functions and pathways, in particular, molecular regulatory activities (regulation of TF activity, phosphorylation), and neural functions such as synaptic activity, gliogenesis, and neural cell differentiation (**Supplementary Table S15**). We also generated a subset of rescued genes that overlapped between any two of CRISPR-, ASO880- and ASO131-rescued genes (**Supplementary Figure S7**). Functional terms derived from these genes (n=410) revealed an enrichment for genes related to axon guidance (p=3.08e-3), a pathway previously identified in our exploratory transcriptomic profiling of XDP neural cells (Aneichyk et al.)^22^, as well as for genes related to regulation of protein ubiquitination (p=3.70e-3), a biological process that we have uncovered to be deranged in XDP postmortem brains (manuscript in preparation). These findings indicate that rescued genes distribute in ontologies that are of relevance to XDP biology, as evident from patient-derived cell lines and tissues.

Finally, we explored potential rescued interactions by constructing a protein-protein network from STRINGdb^75^ using XDP-signature genes (n=102) that belong to the six pathways rescued by CRISPR, ASO131, and ASO880 (**Figure 5C**). This network revealed significant protein-protein interactions (*p<1*.*0e-16*) consisting of 82 connected nodes and 194 edges. Further, the network was enriched for genes localized to the presynapse and for genes with gliogenesis-related functions, as well as DNA-binding transcription factors and their regulators. Highly connected hub genes in this network include *CTNNB1* (Catenin Beta 1) and *HDAC1* (3-hydroxyacyl-CoA dehydratase 1), genes involved in regulation by TF binding and control of neurodevelopment. We also identified interconnected brain-relevant XDP signature genes that are rescued by genome editing and top ASOs, including *GABRA5*, which codes for a subunit of the inhibitory GABA-A receptor in the brain, *PENK* (proenkephalin), and *BAIAP3* (brain-angiogenesis inhibitor associated protein 3). This highlights that our in vitro cell model-based targeting platform can identify tractable signatures that are related to neuronal cell function and maintenance in the human brain (**Figure 5**).

**Figure 5.**
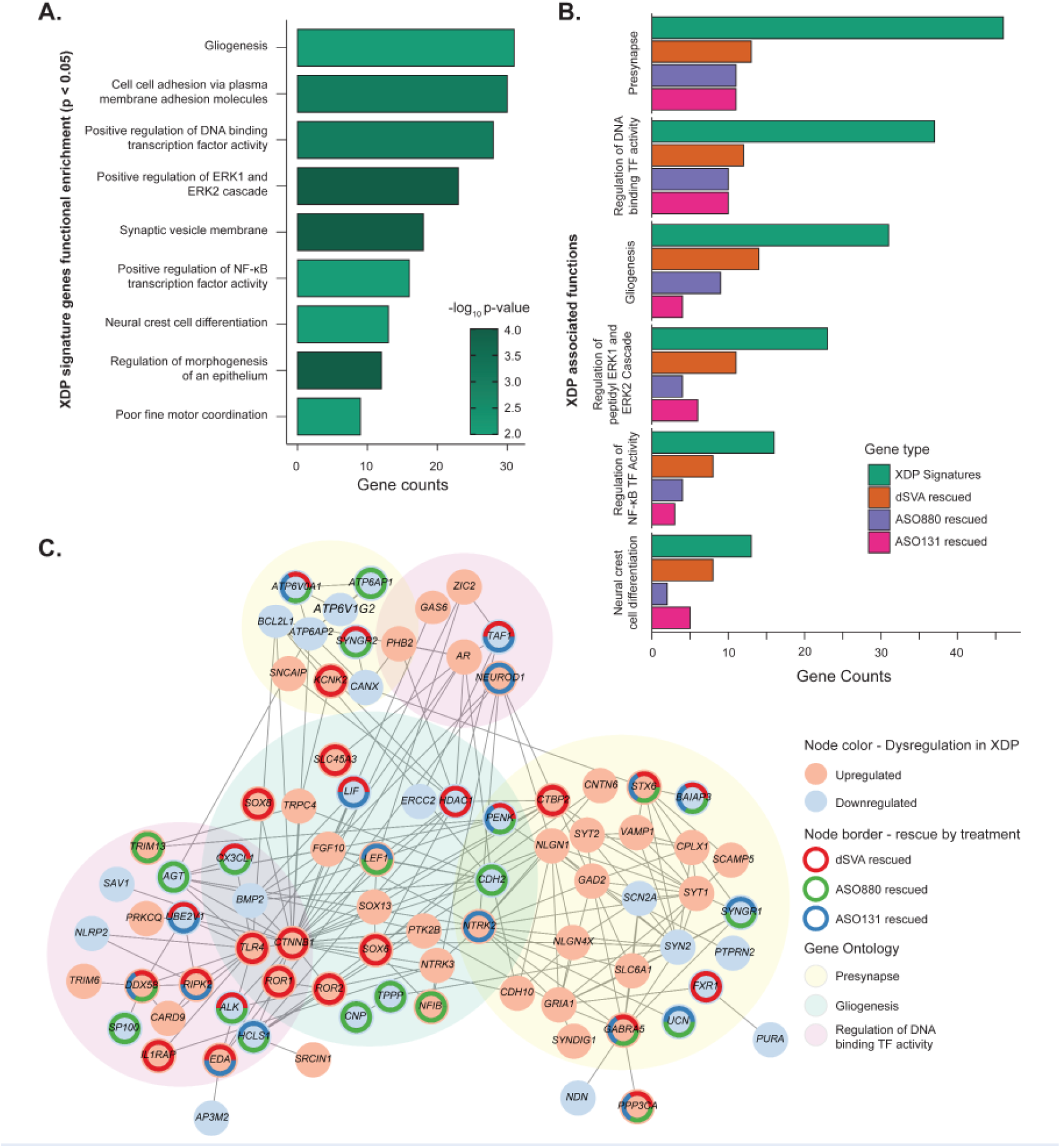
Rescue of dysregulated pathways in XDP NSCs by genome editing and ASOs. **A)** Top significant functional enrichments in XDP-signature genes (p-value < 0.05). **B)** Common significant functional enrichments (FDR < 0.05) in XDP-signature genes rescued by CRISPR, ASO880, and ASO131. **C)** Protein-protein network constructed from genes belonging to functional groups associated with XDP. Overall network enrichment is at p-value < 2e-16. Highly connected nodes include genes rescued by CRISPR, ASOs, or both, as well as functional relevance to XDP phenotype.

Collectively, these analyses suggest that there is a set of convergent genes and pathways that are significantly rescued by CRISPR excision of the SVA and modulation via ASOs. These results may indicate that the collective effects of targeting amelioration of the aberrant expression signatures in *TAF1* have a significant and quantifiable downstream impact on XDP-associated genes, pathways, and regulatory networks.

## DISCUSSION

These studies of XDP offer a window into the fundamental challenges and the future of genomic medicine and precision therapeutics for some classes of rare Mendelian disorders. We previously demonstrated that expression changes in *TAF1* occur in XDP patient-derived neuronal models and are associated with the causal SVA insertion in this gene. *TAF1* is ubiquitously expressed and plays essential roles in the global transcription machinery. Consistent with these properties, in the profiling of global genetic variation from the genome aggregation database (gnomAD), *TAF1* is highly intolerant to loss-of-function variation in males.^30,74,76–78^ In addition to XDP, altered expression of TAF1 has been described as a downstream effect of a causal mutation in other neurodegenerative diseases, such as Huntington’s Disease^79^ and Alzheimer’s Disease.^80,81^ Expression of *TAF1* transcripts has been shown to alter the development of cholinergic neurons, a cell type implicated in dystonic disorders.^70^ Indeed, consideration of the allelic architecture and impact of noncoding mutations is instructive here. In XDP, a noncoding SVA leads to aberrant splicing and modest reduction of *TAF1* expression yet represents a fully penetrant and lethal mutation leading to a late adult-onset cluster of neurodegenerative symptoms. By contrast, damaging missense variants in the same gene are associated with early neurodevelopmental deficits and altered brain morphology in humans^82,83^and model animals.^84–86^ Recent studies have uncovered the role of alternative splicing in gene regulation in the nervous system and its contribution to the molecular pathology of many neurodegenerative disorders,^87^ and *TAF1* AS-i32 itself has been proposed as a reliable marker for monitoring disease^88^. Despite these advances, there was no clear path toward therapeutic amelioration for this debilitating disease. Deciphering the molecular alterations in brain tissue and neuronal cells associated with XDP, alongside the profiling of the functional impact of targeted therapeutic modalities, holds promise for bridging this critical barrier between fundamental biological discovery and relatively rapid therapeutic intervention.

We sought to deconvolute XDP mutational impact from downstream disease signatures to advance precision therapeutics by leveraging a CRISPR/Cas9 engineering approach to excise the causal mutation, correct aberrant splicing and restore *TAF1* expression. We then compared these findings to RNA-based modulation of the splicing abnormality without the need for germline editing in cells. Genome editing reproducibly ameliorated the functional signatures of XDP in patient-derived neuronal cell lines. However, in vivo delivery remains a major challenge that is being actively pursued by the field, with the first in vivo CRISPR therapy in humans recently published.^5^ This barrier is less daunting for ASOs as they readily offer a path to therapeutic delivery via intrathecal injection and have been repeatedly shown to be efficacious in efficiently correcting splicing defects in neurological disorders.^43,89–91^ Our study demonstrates that ASOs are able to significantly repress RNA splicing defects resulting from this SVA insertion, comparable to the effects of CRISPR editing, and have similar degrees of impact on XDP functional alterations at the level of individual genes and pathways. Interestingly, ASOs induce only minimal changes in the overall expression of *TAF1*. Thus, while the rescue of transcriptome-wide signatures suggests that the downstream effects of DNA and RNA-based targeting are equivalent, they propose a question as to whether ablation of the alternative splicing defect is inherently coupled to the restoration of *TAF1* expression, and this remains an area of active investigation and development.

Our analyses revealed an enrichment of dysregulated genes – and their rescue via CRISPR and ASOs – in XDP that are critically important to neurological function and development. These findings reflect transcriptomic changes in the early developmental stage uniquely captured in the NSC lines we used in our testing and screening platform. These include regulators of neural differentiation from stem cell precursors and genes that have been implicated in glial differentiation and activation, such as *HDAC1, CTNNB1, PENK*, and *CX3CL1*.^92–94^ While XDP is an adult-onset disorder characterized by neuronal cell death in its late stages, studies on postmortem brain tissue document neuroinflammation and a prominent glial response in non-atrophic cortical XDP tissue,^95^ similar to what has been described in other neurodegenerative diseases.^96^ Besides gliogenesis-related genes, we also observed rescue of genes related to the synapse and synaptic activity. Rescued XDP signature genes such as *GABRA5, STX6*, and *ATP6V0A1* code for subunits of channels or receptors that map to glutamatergic or GABAergic synapses,^97–99^ cellular components that are relevant for neurotransmission and the movement disorder phenotype observed in XDP. A major limitation in rare neurodegenerative disease research is the scarcity of tissue available for investigation, or the poor translation potential of signatures obtained from diseased and already dead cells. This study highlights the potential of *in vitro* neural cell platforms for scalable translational studies, demonstrating that we can identify relevant genes and pathways that serve as early targets for intervention and biomarker development^100^.

There is emerging evidence to suggest that our approach to ASO screening and functional genomics profiling in neuronal cells offers a path toward a bespoke therapy for XDP. RNA-based strategies have garnered attention due to their therapeutic potential for diseases with splicing defects. There are currently four FDA-approved ASOs for the treatment of Duchenne muscular dystrophy (DMD), all of which promote exon skipping to restore the open reading frame of dystrophin.^9,101–103^ The FDA-approved drug milasen was designed to block a cryptic splice site in Batten disease,43 while ASO-directed inclusion of exon 7 in the alternatively spliced *SMN2* gene is the mechanism of action of the FDA-approved ASO nusinersen for the treatment of spinal muscular atrophy (SMA).^104^ In preclinical studies, ASO-mediated skipping of *LRRK2* exon 41 has been shown to restore mitochondrial function in Parkinson’s Disease (PD)-derived fibroblasts and endoplasmic reticulum calcium regulation in iPSC-derived neurons.^105,106^ ASO-targeted removal of exon 17 in APP, which contains the γ-secretase site responsible for amyloid cleavage, has led to decreased Aβ42 expression *in vitro* and *in vivo* and is currently being investigated as a future therapy for Alzheimer’s Disease dementia.^107^ Given the acute need to identify novel and tractable targets and treatment approaches for this lethal disorder, our strategy using in vitro neuronal models provides a significant leap forward in understanding the cellular processes that are disrupted in XDP. These data now set the stage for clinical studies into gene-based therapies for this devastating Mendelian disorder.

## Data Availability

Data will be added to dbGAP study phs001525.v2.p1 upon publication of the article

## ACKNOWLEDGEMENTS

This research was supported by the Collaborative Center for X-Linked Dystonia Parkinsonism (CCXDP) at the Massachusetts General Hospital, the National Institutes of Health (R01NS102423, R00NS118109, U01HG011755, R01MH115957), and the Massachusetts General Hospital Executive Committee on Research (ECOR) Fund for Medical Discovery.

## AUTHOR CONTRIBUTIONS

Conceptualization, R.Y., C.A.V., A.D., S.R., D.C.B., and M.E.T.; Methodology, R.Y., C.A.V., A.D., S.R., S.S. D.G.; Investigation, R.Y., C.A.V., A.D., S.R., S.S., D.G., S.E., K.O., M.Sa., J.L., R.B., M.A.M., M.J., M.C., M.G.M., C.F.-C., G.P.A.L., M.Sy., M.S.V.-A., E.L.M., M.A.C.A., and C.C.E.D.; Formal Analysis, R.Y., C.A.V., A.D., S.R., D.G., and Resources, M.A.M., M.J., M.C., C.F.B., E.B.P. and L.O.; Visualization, R.Y., C.A.V., A.D., and S.R.; Writing – Original Draft, R.Y., C.A.V., A.D., and S.R.; Writing – Review & Editing, all authors; Supervision, L.O., N.S., C.F.B., D.C.B., and M.E.T.; Funding Acquisition, D.C.B., and M.E.T.

## MATERIALS AND METHODS

### CRISPR editing of XDP-specific SVA in iPSCs

Clinical characterization, collection, and reprogramming of cell lines used in this study have been previously described.^22,71^ CRISPR editing to remove the XDP-specific SVA in iPSCs was modified from prior XDP genome editing experiments^22,49^ to achieve higher targeting efficiency. Specifically, iPSCs were electroporated in suspension with a high performance Cas9 variant (TrueCut Cas9 Protein V2; Thermo) and guide RNAs (gRNA 5: ACATATACCATTCTATTCTT and gRNA 8: GATGTGGAAAAAAAATGTAC) using the Neon Transfection System and the following settings: 1400 V, 20 ms, 1 pulse. Transfected iPSCs were plated on Geltrex-coated 24-well plates in StemFlex media without antibiotics and 10 μM rock inhibitor. Once confluent, genomic DNA was isolated, and PCR was performed to identify SVA excision. Samples with suspected SVA excision were subsequently sent for Sanger sequencing to confirm targeting. Successfully edited iPSC pools were collected as single cells and plated at 10K or 20K per 10 cm dish for clonal selection in StemFlex media containing 10 μM rock inhibitor. Media was exchanged 48 hrs post-plating and every other day thereafter until colonies reached 2-4 mm in diameter, approximately 7-10 days after plating. Single colonies were manually picked into individual wells of a 96-well plate until wells reached 90% confluency. Each clone was then collected with diluted accutase (1:3 in PBS), and plated into 3 replicate 96-well plates: one to which freezing media (80% FBS/20%DMSO) was added 1:1 to each well and stored at -80°C as a clone recovery plate; two coated with geltrex to generate both a passaging and a genomic DNA plate. Media was exchanged on the culture plates every other day. Once cells reached 90% confluency, genomic DNA was collected from one of the sister plates for clonal screening via PCR. Editing resulted in more than 150 iPSC clones in which the SVA was excised. From that pool, 3 to 5 clones per parent XDP line with no other insertions/deletions in the targeted region were expanded. A select number of targeted lines from each parent XDP line were sent for karyotyping, and each was shown to have a normal karyotype. Lines exposed to CRISPR/Cas9 machinery but retained the SVA (unedited XDPs) were treated as the naïve XDP lines (those not exposed to targeting) in subsequent transcriptomics analyses, as described in the results.

### Post-mortem brain tissue

Post-mortem control brain tissue was obtained from the NIH NeuroBioBank. Port-mortem XDP brain tissue was obtained from the Collaborative Center for XDP Brain Bank at Massachusetts General Hospital (Boston, MA USA). These procedures were approved by Institutional Review Boards at Makati Medical Center (MMCIRB 2017-134) and Massachusetts General Hospital (protocol 2016P000 Sharma). The methodology of brain donor identification, consent, tissue extraction and tissue processing has previously been reported^59^. In brief, half of each brain was processed for fresh frozen tissue. Each hemisphere was cut into 1 cm thick sections and frozen between Teflon-coated steel plates in dry ice and then stored at -80°C. Regions of interest were then identified and dissected from the frozen sections while on dry ice. RNA extraction was performed from ∼50 mg of tissue using Trizol and standard kits (QIAgen).

### ASO Design

ASOs were designed and modified by Ionis Pharmaceuticals. 20-mer antisense oligonucleotides with uniform 2′-O-methoxyethyl (MOE) chemistry were designed to cover approximately 100 nt +/-the exon32/ intron32 junction and the intron32/*TAF1* AS-i32 junction. Top 80 oligos were selected based on low off-target hits in the human transcriptome and separated by at least two nucleotides. Second generation ASO design, based on the target sequences of previous top hits, included additional proprietary base modifications to increase stability, target engagement and neuronal uptake.

### ASO treatments

Fibroblast ASO transfection: Fibroblasts from a single patient line (MIN#34427) were plated in fibroblast growth medium (DMEM containing 20% FBS and 1% Pen/Strep) at 6K/well in 96 well plates in duplicate one day before transfection. The following day, ASOs were transfected at 100 nM final concentration using Cytofectin (Genlantis, San Diego, CA) according to manufacturer’s instructions. Briefly, 1 μL of cytofectin reagent was added to 1 mL of Opti-MEM in a single well of a deep well 96 well plate. ASO (5 μL of 20 μM) was added to cytofectin:Opti-MEM and manually mixed by pipetting. Media was removed and cells were washed with 1X PBS. A total of 100 μL of ASO:cytofectin mixture was added to cells and incubated at 37°C. After 4 hours, the transfection solution was removed and replaced with fresh growth medium. RNA was isolated 48 hours after transfection using Trizol.

#### Dose Response Curve Analysis (NPCs)

Neural progenitor cells (NPCs) were generated as previously described.^66,108^ NPCs were plated in 96-well plates at 20K/well in duplicate in neural progenitor media (NPM; DMEM/F-12 containing 2% B-27, 1% Pen/Strep, 2 μg/ml Heparin, 20 ng/ mL bFGF and 20 ng/mL EGF) media at day 0 (D0). Media was changed on D1 with fresh NPM containing 0, 0.03, 0.1, 0.3, 1, 3, 10, or 30 μM ASO. Cells were lysed on D4 in Trizol and stored at -80°C until RNA isolation. RNA isolation was performed using Direct-zol 96 RNA kits (Zymo, cat# R2054).

#### NPC ASO treatments for RNA Capture-Sequencing

A total of 9 XDP, 2 ΔSVA (generated from original targeting performed^22^ and 5 control NPC clones were plated in 12-well plates at 100-120K cells/well in NPM media on D0. The media was exchanged on D1 and D4 with fresh NPM containing 5 μM ASO or equivalent volume of H2O. Cells were lysed on D7 and stored in Trizol at -80°C until RNA isolation. RNA isolation was performed using Direct-zol RNA micro columns (Zymo, cat# R2062).

#### NSC ASO treatments

Neural stem cells (NSCs) were generated as previously described.^22^ NSCs were thawed at passage 2. When cells were confluent, NSCs at passage 3 were plated into duplicate 24-well plates at 60K/well in neural expansion media (NEM; 50% Neurobasal medium, 50% Advanced DMEM/F-12, 2% Neural induction supplement and 1% Pen/Strep) containing 1:2000 rock inhibitor on D0. The media was exchanged on D1 and D4 with fresh NEM containing 5 μM ASO or the equivalent volume of H2O. Treatments were organized into three batches, with each batch containing a collection of each genotype along with the naïve proband XDP line. This generated repeat reads of each of the targeted probands, bringing the sample number up to 71 per candidate ASO. Cells were lysed on D7 in 100-140 μl RLT buffer and frozen at -80°C. For RNA isolation, the total volume of RLT per sample was transferred to a 2 mL costar plate and additional RLT was added for 250 μl total volume. Trizol LS (750 μl) was added and samples were mixed and incubated at RT for 5 min. Chloroform (200 μl) was added, then incubated at RT for 2 min. Samples were then spun at 3000 rpm for 5 min at RT. A total of 420 μl of aqueous phase was transferred to a 1 mL costar plate and an equal volume of 70% ethanol was added. Samples were mixed, then transferred to a Qiagen RNeasy 96 well plate and vacuumed according to protocol. Samples were washed with RW1 buffer and incubated with 100 U DNase I per well in 1X DNase buffer for 15 min. Samples were washed once with RW1 buffer, then twice with RPE buffer. Samples were spun down at 5000 rpm for 10 min. RNA was eluted in 100 μl water into a new 96-well plate. RNA was further concentrated down to 20-25 μl using a Genevac evaporator at 27°C for 30 min. RNA integrity was spot-checked per plate to ensure quality.

### AS-i32 detection via real time PCR

To amplify AS-i32 for initial screening in fibroblasts and NPC dose-response curve analysis, cDNA synthesis was performed on 20-150 ng input RNA using Superscript III or IV (Thermo Fisher) with Oligo DT primers. Synthesized cDNA was then used as a template for 10-15 cycles of PCR to amplify the *TAF1*-32i target region, as previously described.^66^ Preamplification product was concentrated using the Zymo Clean and Concentrator Kit (Zymo, cat# D4004) down to 15 μl (Fibroblasts) or 20 μl (NPCs). A total of 1 or 2 μl of pre-amplified product was used per real time reaction. The 2^(-ΔCT)^ method was used to calculate relative expression levels of AS-i32. For fibroblasts, the housekeeping gene *GAPDH* was used. For NPCs, the housekeeping gene *GUSB* was used. Relative levels were then normalized to no ASO levels (untransfected or media only controls) to determine efficacy of ASO treatments. To calculate EC_50_s, fit curves were calculated using nonlinear fit (log(inhibitor) vs. response - variable slope) and constraining the top and bottom values at 1 and 0, respectively, using GraphPad Prism 10 software.

### Transcriptional profiling of ASO treatments in NSCs

#### Strand-specific dUTP RNAseq library preparation

RNASeq libraries (n=469) were prepared using TruSeq® Stranded mRNA Library Kit (Illumina) and prepared per manufacturer’s instructions. In brief, RNA sample quality (based on RNA Integrity Number, RIN) and quantity were determined on an Agilent 2200 TapeStation and between 500-100 ng of total RNA was used to prepare libraries. 1 μL of diluted (1:100) External RNA Controls Consortium (ERCC) RNA Spike-In Mix (Thermo Fisher) was added to each sample alternating between mix 1 and mix 2 for each well in batch. PolyA bead capture was used to enrich for mRNA, followed by stranded reverse transcription and chemical shearing to make appropriate stranded cDNA inserts for the library. Libraries were finished by adding both sample-specific barcodes and adapters for Illumina sequencing followed by between 10-15 rounds of PCR amplification. Final concentration and size distribution of libraries were evaluated by 2200 TapeStation and/or qPCR, using Library Quantification Kit (KK4854, Kapa Biosystems), and multiplexed by pooling equimolar amounts of each library prior to sequencing. RNASeq libraries were sequenced to 30-40 million reads per library on Illumina NovaSeq machines.

#### RNA Capture-Sequencing (CapSeq) library preparation

RNA CapSeq libraries (n=1145) were prepared using a combination of TruSeq® Stranded mRNA Library Kit (Illumina) and Agilent SureSelectXT HS2 kit with a custom capture library targeting the region of 400 kb on the X chromosome from *OGT* to *CXCR3*. cDNA was made from RNA using the TruSeq® Stranded mRNA Library Kit (Illumina). 500-100 ng of total RNA was used to prepare libraries that were first bead captured to enrich for mRNA, followed by stranded reverse transcription to generate appropriate stranded ∼175 bp length cDNA inserts. These cDNA inserts were end-repaired, adenylated, ligated to adapter oligos and then amplified with 5 cycles of PCR according to manufacturer’s instructions. After quantification, ∼90 ng of each amplified library was hybridized overnight with the capture baits. Following capture cleanup, each pooled library was amplified with additional 16 cycles of PCR. Final products were quantified using the TapeStation 2200 and pooled for rapid mode sequencing. RNA CapSeq libraries were sequenced at the Broad Institute Genomic Services as 150 bp paired-end reads on an Illumina Novaseq platform.

### RNAseq and RNACapSeq data-analysis

RNASeq preprocessing: FastQC was used to determine the data quality of the sequencing data. Read pairs of RNA CapSeq and RNAseq were trimmed using trimmomatic v0.36 for Illumina Truseq adapters and primers. The trimmed read pairs were then aligned to human genome (GRCh37, Ensembl release 75) with SVA inserted at position X:70660363 by STAR 2.5.2b allowing 5% mismatches with a unique mapping and with following parameters “--outFilterMultimapNmax 1 --outFilterMismatchNoverLmax 0.05 --alignEndsType EndToEnd”. Counts were generated using STAR (v2.5.3a) with the following non-default arguments --outFilterMultimapNmax 1 –outFilterMismatchNoverLmax 0.05 --alignIntronMin 21 --alignIntronMax 0 --alignEndsType EndToEnd --quantMode GeneCounts --twopassMode Basic. Relative normalization factors among samples were calculated by applying DESeq2 function estimateSizeFactor() on the count matrix of each gene across all samples.

### De novo transcript assembly and quantification

For de novo assembly, alignments of each sample (both total RNAseq and RNA CapSeq) within the *TAF1* genomic region were deduplicated and reads pairs from the same brain regions (for postmortem brain) or genotype (for NSCs) were then merged. Transcripts were assembled using Trinity v2.2.0 with the following parameters: default settings “--SS_lib_type RF”, “--genome_guided_bam” and “--genome_guided_max_intron 100000”.^109^ After the preliminary assembly, transcripts were compared and merged into a non-redundant list using custom scripts. These transcripts were then compared against the human genome GRCh37.75 using BLAT to resolve internal splicing structures and for annotation. For each transcript to be included in the final reference index, we required the transcript to be identified in 40% of RNAseq samples within the group (i.e., brain region or genotype); else, the de novo transcript was disregarded. After the index was built for each group, RSEM was applied to quantify transcript variant usage as a percentage of overall *TAF1*. Expression of the transcript was then computed for using this percentage and the corrected, normalized expression of *TAF1* as above.

### Differential gene expression analysis in NSCs between XDP and Control samples

For our NSC samples we have two sets of samples derived from XDP patients: *naive* XDPs (n=9), which were never exposed to CRISPR/ Cas9 or gRNA targeting the SVA, and unedited XDPs (n=21), which were exposed to CRISPR/Cas9 and gRNA but did not have the SVA excised from the genome. After confirming that the gene expression profiles of these sets of samples were highly correlated (**Supplemental Figure S1A-E**), we pooled these samples together to form a single set of XDP samples (n=33). Two of these 33 samples were outliers as they were > 2 standard deviations from the centroid of samples in PCA space and thus excluded from all subsequent analysis. Within the text, when we refer to XDP NSC samples, we refer to this pooled group of 31 samples, unless otherwise specified.

We use a Generalized Linear Model (GLM) based approach to estimate differential expression effect size and significance between untreated XDPs vs untreated Controls. The first step is filtering out genes with low expression (CPM < 0.1) in at least 50% of either genotype (Controls or XDPs). Next, we used the DESeq2 package to normalize the raw RNAseq counts by library size for the retained genes.^110^ The sva package^111^ was used to estimate Surrogate Variables (SVs) from the size-normalized counts to correct for known cofactors like batch as well as unknown technical factors. For each gene, we fit a GLM using the gene’s normalized expression and all estimated SVs as predictor covariates to estimate an effect size (log fold-change) and significance (p-value, FDR) for that gene’s expression on predicting the sample group (e.g. genotype) (**Supplementary Table S12**).

### Robust statistical modeling of gene expression changes in NSCs

We also applied a bootstrapping approach to identify differential expression patterns between untreated XDPs and untreated controls to compensate for the unbalanced sample sizes. To compute an individual bootstrap, we first select a random set of 12 samples, 6 untreated controls and 6 untreated XDP, and apply the differential gene expression workflow specified above (CPM filtering, size normalization, SV estimation, GLM fitting and multiple-testing correction) to estimate effect size and significance for all genes for this set of 12 samples. We computed these results for 52,800 bootstraps (i.e. 52,800 random sets of 12 samples each), giving us distributions of logFC and p-values/FDR for each gene across all bootstraps. 16723 genes pass the CPM cut-off in all bootstraps, and 21903 genes are tested in at least 1 bootstrap. From this, we can calculate the frequency across all bootstraps that a gene 1) passes the CPM thresholding 2) is up/down-regulated (log fold-change > 0, log fold-change < 0, respectively) 3) is up/down-regulated with a specific statistical significance (p-value < 0.05 or FDR < 0.1) (**Supplementary Table S7**). FDR correction is done across all genes within each bootstrap, but separately for each bootstrap.

### Differential gene expression analysis in NSCs using paired treated and untreated XDP samples

Each untreated XDP sample has paired samples after each ASO treatment (7 ASOs) and the CRISPR/Cas9 editing, profiled using RNAseq. We made use of this sample pairing when calculating the effect size of each treatment DEGs to account for individual genetic variation between samples and compared untreated XDP vs untreated control samples as estimated by the GLM approach outlined above. The CPM filtered size-normalized gene counts were adjusted by SVs and log transformed. The corrected counts were used to estimate gene expression for each sample for each gene. Specifically, we calculate the (gene x sample) matrix of “adjusted counts” *Y*_*j*_ for a given treatment *j* as follows

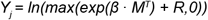

with β being the (gene x variable) matrix of GLM coefficients, *M* being the (sample x variable) model matrix indicating the estimated SVs and Genotype of each sample, and *R* being the (gene x sample) matrix of GLM residuals. Here, the variables are all the input variables to each GLM model i.e. Intercept and all estimated SVs. So, for each treatment *j* we separately compute the matrix *Y*_*j*_, comparing untreated XDPs and treated XDPs, giving us 8 separate (gene x sample) matrices of adjusted read counts. The adjusted counts are >99% correlated across treatments for the untreated XDP samples (**Supplementary S8**). Using these adjusted counts, we computed the effect size for a gene g for each pair *i* of treated/ untreated samples (*t*_*i*_,*u*_*i*_) as follows

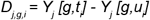

since *Y*_*j*_ is on the log scale this difference represents a log fold-change for each pair of samples. So *D*_*j,g,i*_ > 0 implies sample *i* shows increased expression of gene *g* when treated with treatment *j* and vice-versa. Within each treatment, for each gene we calculate *D*_*j,g*_, the mean log fold-change over all sample pairs and a corresponding 95% confidence interval. All *D*_*j,g*_ values and 95% confidence intervals are available as a supplementary table (**Supplementary Table S8**).

### Intron retention analysis

IRFinder from Middleton et al. was applied to estimate genome-wide IR in each sample.^112^ In brief, the IR level of each intron is estimated by the package as a ratio of the reads in intronic regions to reads splicing across flanking exons. To assess the effect of XDP genotype and ASO treatments on IR ratio, a general linear model (GLM) was fitted to correlate IR ratio (via link function of identity) with genotypes and treatments. See “Differential gene expression analysis in NSCs using paired samples” session for model fitting details and differential IR analyses.

### Gene rescue criteria

We use two criteria to classify genes as “rescued” or not by the treatments (ASO or CRISPR editing). For the “nominal rescue criteria” a gene is considered rescued by a given treatment if 1) the gene is an XDP signature gene and 2) the GLM log-fold-change comparing treated XDPs to untreated Controls is not significantly different from 0 (z-test p-value > 0.05). All genes and their rescue classifications are provided in **Supplementary Table S14**.

### Functional enrichment analysis

We tested the functional enrichment of various groups of genes (e.g. DEGs, co-expression modules, rescued genes, etc.) using one-tailed Fisher’s exact test for the curated lists of Gene Ontology (GO) Terms, Canonical Pathways, Human Phenotype Ontology (HPO) terms, and Transcription Factor Targets (TFT) from MSigDB (v2022.1.Hs).^113^ We also included a list of TAF1 targets^114^ and a list of genes from the literature with functional associations to neurodegenerative phenotypes. The background set of genes for each comparison was the intersection of all genes annotated in a given ontology and all genes considered in each analysis.

