## Supplementary Info for "Therapeutic targeting of alternative splicing caused by a lethal noncoding structural variant in X-linked dystonia parkinsonism"

***Supplementary information***

#### Supplementary Tables

**Table S1.** Description of samples used in the transcriptomics study.
**Table S2.** AS-i32 in NSC lines using deep transcript sequencing (RNA-CapSeq)
**Table S3.** qPCR detection of AS-i32 and GAPDH from fibroblasts ASO transfections. Table shows replicate reads and normalization calculations.
**Table S4.** qPCR detection of AS-i32 and GUSB from original 12 ASO treatments in 2 NPC lines. Table shows replicate reads and normalization calculations.
**Table S5.** qPCR detection of AS-i32 and GUSB from second generation ASO treatments in 3 NPC lines. Table shows replicate reads and normalization calculations.
**Table S6.** TAF1 IR-i32 in brains and NSC samples
**Table S7.** The results of the bootstrapping analysis for all genes with the following columns; EnsemblID: the EnsemblID of the gene expression_pattern: which expression pattern the results are for (up, down, nominal, nominal_and_up, nominal_and_down, fdr, fdr_and_up, fdr_and_down) bootstraps: the number of bootstraps where this gene passed the CPM thresholding (max is 52,800 total number of bootstraps computed) raw_value: the number of bootstraps in which gene shows the specified expression_pattern fraction_of_bootstraps: raw_value / bootstraps. The expression patterns are defined as follows; up: the gene has an LFC > 0 down: the gene has an LFC < 0 nominal: the gene has a p-value < 0.05 nominal_and_up: the gene has an LFC > 0 and p-value < 0.5 nominal_and_down: the gene has an LFC < 0 and a p-value < 0.05 fdr: the gene has a BH p-value < 0.1 fdr_and_up: the gene has an LFC > 0 and a BH p-value < 0.1 fdr_and_down: the gene has an LFC < 0 and a BH p-value < 0.1.
**Table S8.** ASO screening results for TAF1 in NSCs comparing paired ASO-Treated/CRISPR-edited XDP samples and untreated XDP samples.
**Table S9.** TAF1 transcripts profiled across brain regions.
**Table S10.** Differential expression results comparing naive XDPs vs Controls**.**
**Table S11.** Differential expression results comparing unedited XDPs vs Controls.
**Table S12.** Differential expression results comparing all XDPs vs Controls.
**Table S13.** Functional enrichment results for the XDP signature set of genes (n=1460).
**Table S14.** Classification of each gene as “Rescued” or “Not Rescued” for each treatment.
**Table S15.** dSVA, ASO880 and ASO131 rescued XDP pathways and genes.

### Supplementary figures

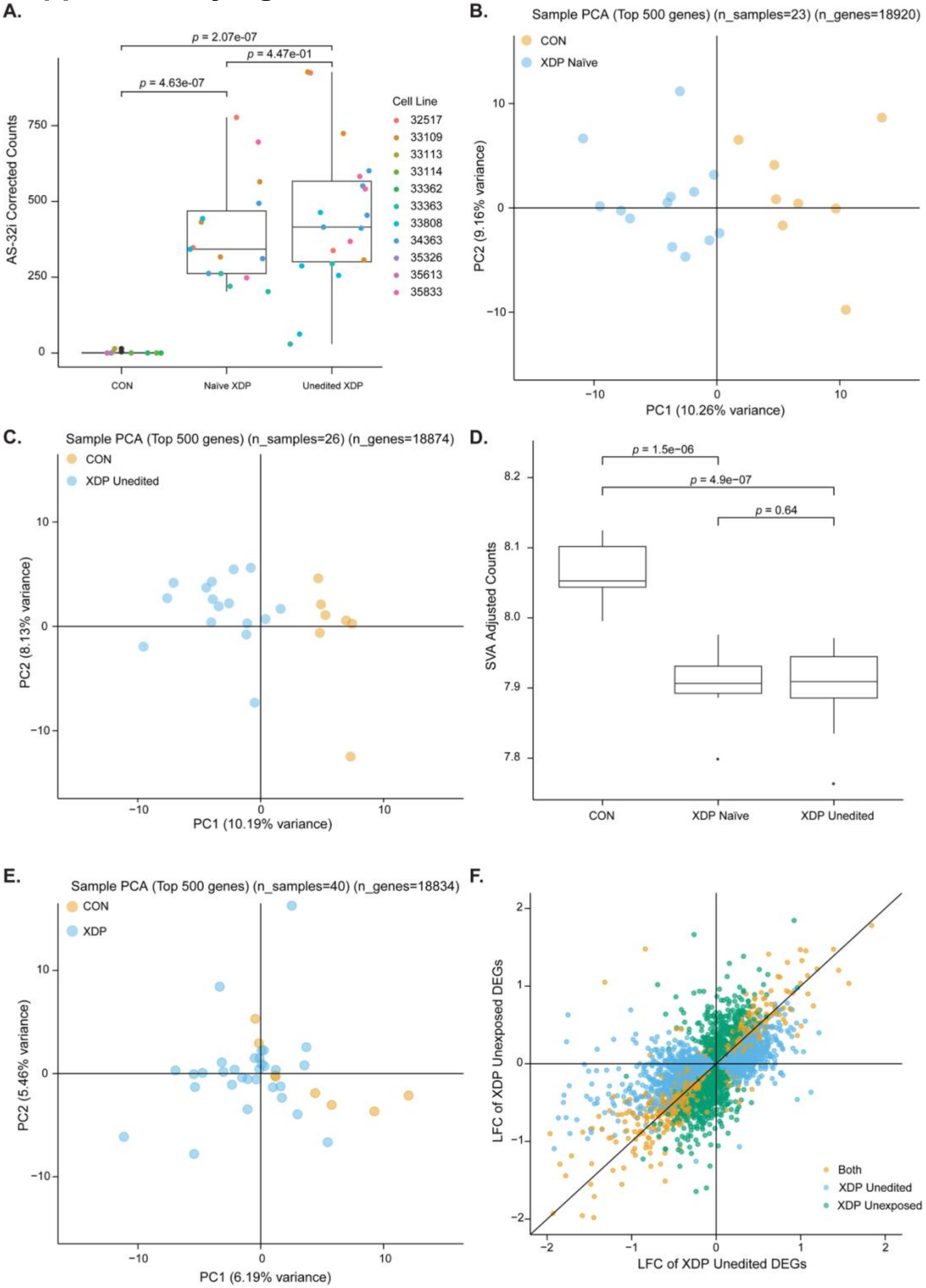


**Figure S1. A)** TAF1 AS-i32 in Controls, naïve XDP and unedited XDP NSC lines. **B)** PCA of Control and unexposed XDP samples computed from the adjusted counts of the 500 most variable genes. **C)** PCA of Control and unedited XDP samples computed from the adjusted counts of the 500 most variable genes. **D)** The x-axis is the LFC of genes when comparing unedited XDP vs Controls. The y-axis is the LFC of genes when comparing naïve XDP vs Controls. Genes are colored based on whether they are DEGs ($p < 0.05$) in either comparison or both. Genes that are not DEGs in either comparison are omitted. **E)** Distribution of adjusted counts for Control, naïve XDP and unedited XDP samples, p-value is of t-test comparing means. **F)** PCA of Control and all XDP samples computed from the adjusted counts of the 500 most variable genes.

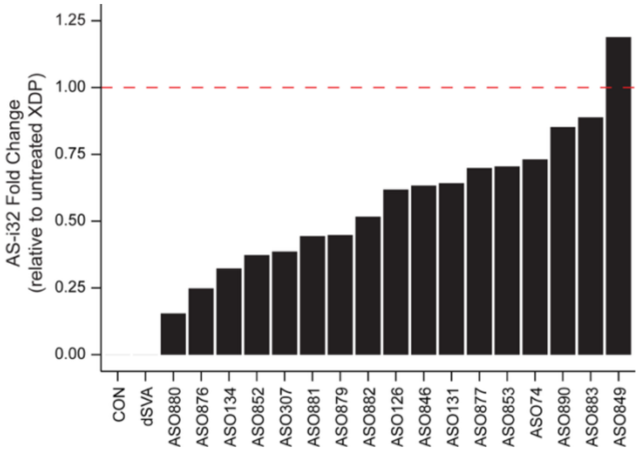


**Figure S2.** Fold change of TAF1 AS-i32 in untreated control, CRISPR XDP (ΔSVA) and ASO-treated XDP NPC clones compared to untreated XDP (red line).

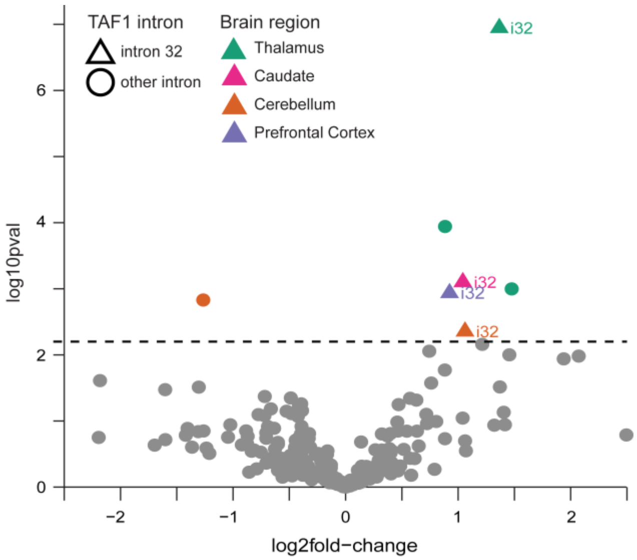


**Figure S3.** Intron retention in all *TAF1* introns across brain regions. When comparing XDP to Controls, only intron 32 (i32) showed significant difference ($p<0.10$) in intron retention for multiple brain regions.

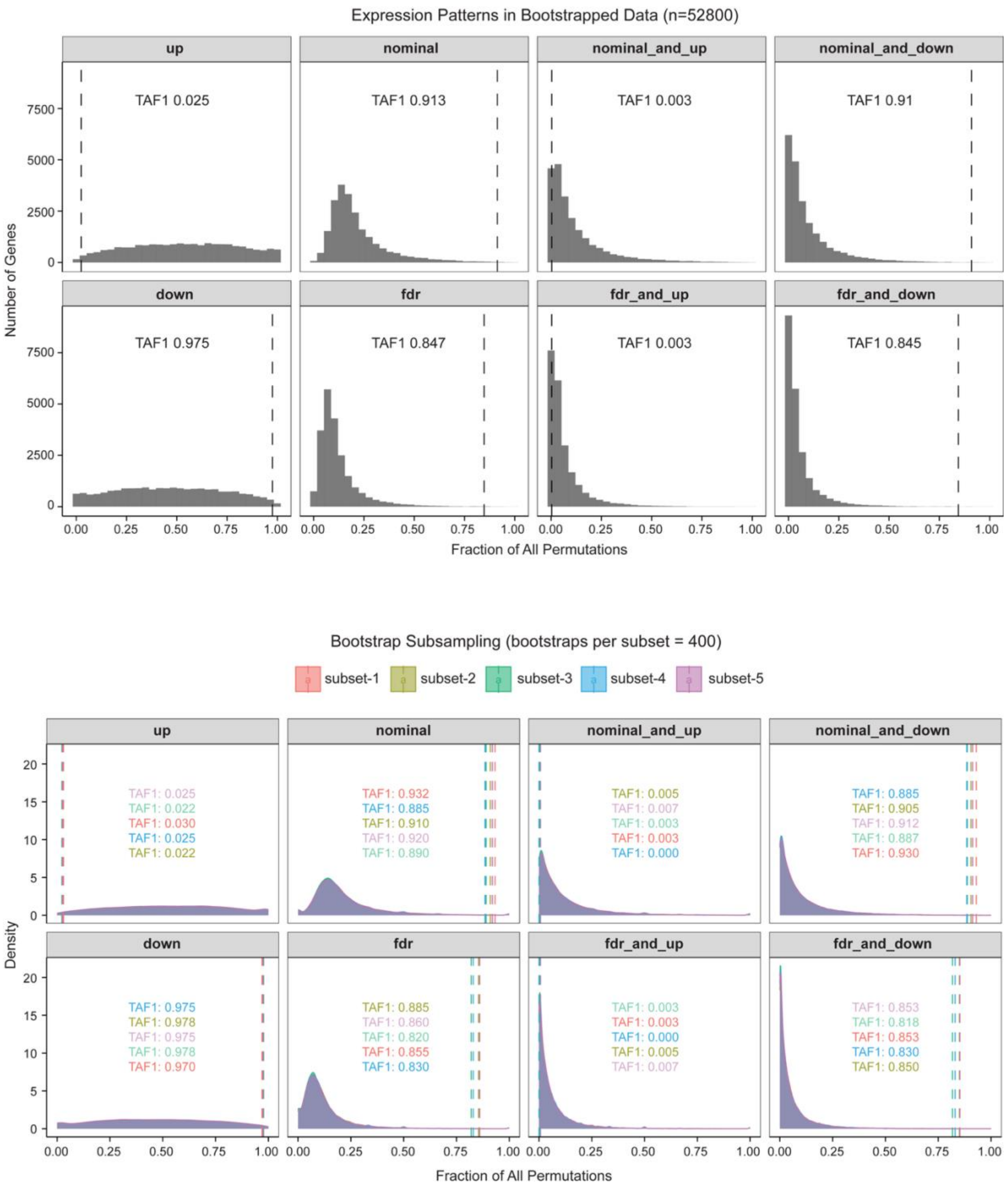


**Figure S4. A)** The frequency of gene expression patterns from the bootstrapping analyses. The x-axis is the frequency that a gene shows the specified expression pattern across all bootstraps where the gene passes the CPM filter. The vertical line represents the frequency at which TAF1 shows the specified expression pattern. Up/Down refer to genes being up/downregulated in XDP relative to Controls respectively. **B)** The same representation of the data as Fig. S4A but plotting the frequency distributions of 5 different random subsets of 400 bootstraps each.

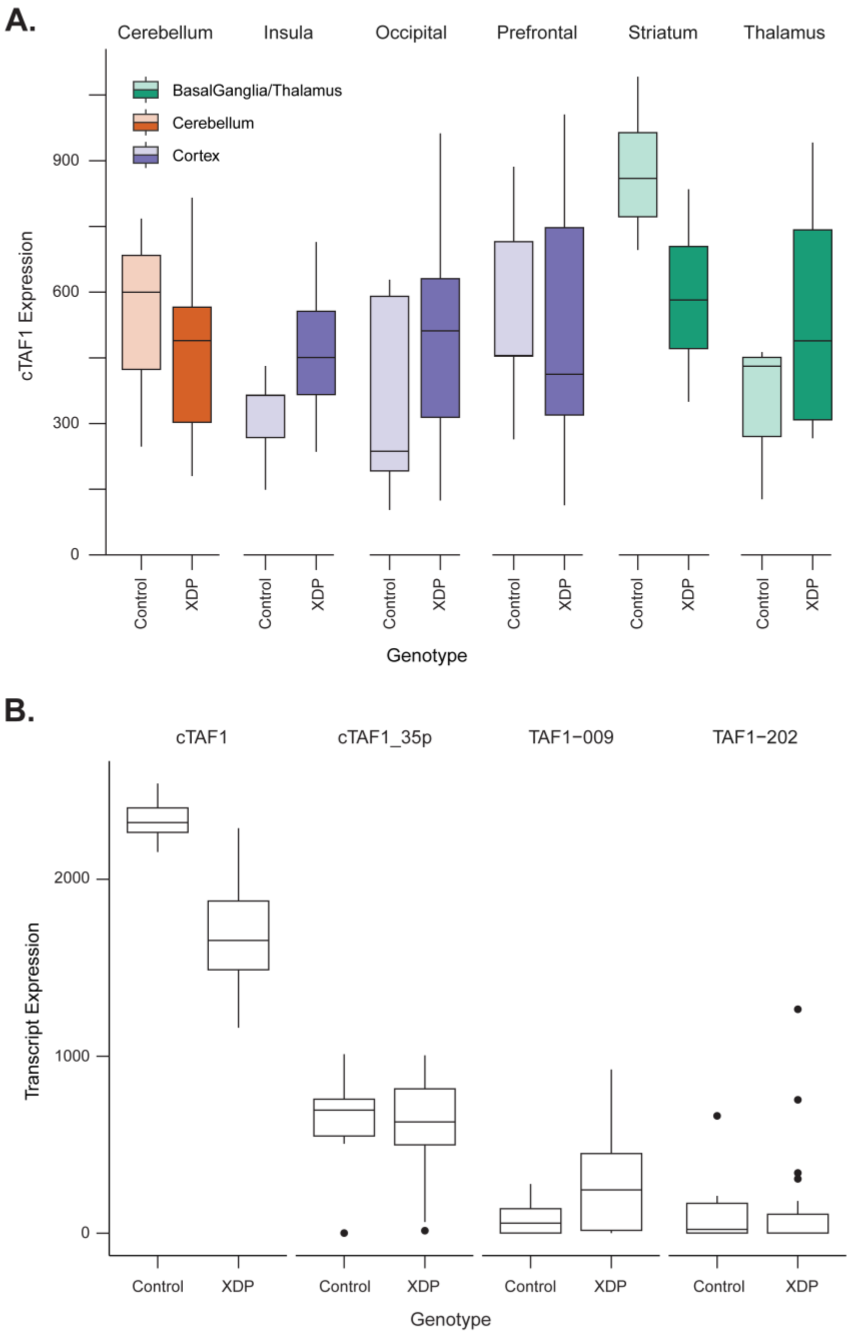


**Figure S5. A)** cTAF expression in XDP and Control samples across brain regions. Significant difference is seen between groups in samples from the caudate (p=0.03). **B)** *TAF1* transcript variants in NSCs. cTAF1 is the predominant transcript variant.

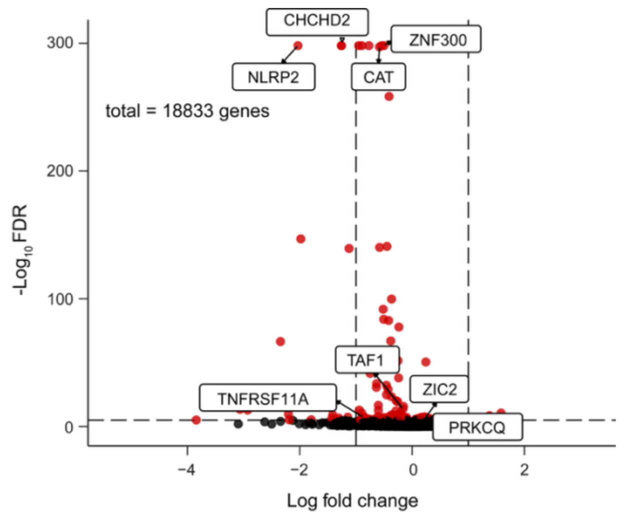


**Figure S6:** Distribution of fold-changes of differentially expressed genes (FDR <0.1) obtained by comparing XDP and Control RNASeq profiles.

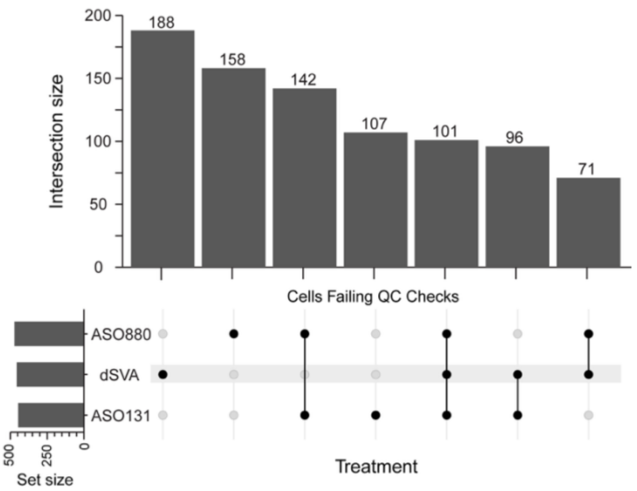


**Figure S7:** Sharing of rescued XDP-signatures by CRISPR-editing of XDP-specific SVA and top 2 ASOs with rescue of *TAF1* AS-i32, IR-i32 and *TAF1* expression.
